# Post menopausal women with Musculoskeletal disorder (MSKDs) and its relationship with regular dietary habit

**DOI:** 10.1101/2025.01.13.24319642

**Authors:** Md Faruqul Islam, Md. Omar Faruque, Md. Alauddin, Md Feroz Kabir, Golam Moula, Akter Hossian, Zahid Bin Sultan, Shahid Afridi, Md Waliul Islam

**Author notes:** **Corresponding author** Shahid Afridi, Department of Physiotherapy, Saic College of Medical Science and Technology (SCMST), Bangladesh.

## Abstract

**Objectives:** to examine the association between consistent dietary patterns and musculoskeletal diseases (MSKDs) in postmenopausal women.

**Methods:** A descriptive cross-sectional study recruited 576 women aged 42 to 70 years who received treatment at the Centre for the Rehabilitation of the Paralyzed (CRP), specifically the Musculo-Skeletal Department, located in Savar, CRP-Dhaka Mirpur Centre, and Unique Pain and Paralysis Centre (UPPC), Dhaka Mirpur, Bangladesh during the period from December 2022 to December 2023. The musculoskeletal disorders (MSKDs) were determined by Nordic MSK Questionnaire (NMQ) and regular dietary habit was determined by the 24-hour Dietary consumption Questionnaire. Data were collected by face-to-face interviews on demographic factors, medical history, menopausal state, and symptoms. Correlation models were used to identify factors associated between MSKDs and dietary habit.

**Results:** Among all participants, the age was categorized in several years and no one experiencing menstrual cycle in last 3 month. The mean overweight and obese BMI were 38.50% and 9.20%. All most all participants were feeling chronic pain on various part of body last 12 months and mean pain rating were 6.111.321. As well as dietary intake were calculation of each individual throughout a 24-hour period. The chronic MSKDs and protein was associated with upper back(p=0.046), carbohydrate was associated with LBP (p=0.005), hip pain (0.008) and knee pain (0.002) while fat was associated with shoulder pain (p=0.007*), wrist pain (p=0.002*), hip pain (p=0.044*), knee pain (p=0.002*), and ankle pain (p=0.010*) but acute pain was only associated with carbohydrate (p=0.012) which had significant.

**Conclusion:** Post menopausal women with acute MSKDs had suchlike no relation with regular dietary habit but a substantial association was seen with some chronic MSKDs.

## Introduction

Menopause is the termination of menstrual cycles and the decline in female hormone synthesis by the ovary. It commonly happens between the ages of 45 and 55 in women^1^. The phrase “postmenopausal period” is used to describe the phase of life that occurs following menopause. During the post menopausal age, there is a dramatic fall in the levels of estrogen and progesterone hormones^2^. Epidemiological studies have revealed that 65-85% of women have difficulties at menopause^3^. Women encounter major physiological changes that may have a dramatic impact on their social lives, emotional well-being, physical ability, and general quality of life^4^.

Musculoskeletal diseases (MSKD) are defined by reduced function, limited activities, and restricted mobility, with gender-based differences^5^. It rates as the second most prominent factor leading to impairment globally, as judged by Years Lived with impairment (YLD). MSK illness causes for roughly 17% of impairment in postmenopausal women^6^. The estimated growth from 1990 to 2010 was 45%^7^. MSKD is linked with dietary behaviors such as vitamin D and calcium consumption, smoking, and alcohol usage^8^. This is generally attributable to a shift in food patterns. Hence, women often have less interest for engaging actively in various hobbies^9^.

Annually, about 6 million women worldwide are experiencing menopause. Annually, about 1.3 million women in the United States undergo menopause^10^. The prevalence of premature menopause in black and Hispanic women is 3.7% to 4.1%, while it is 2.9% for white women^11^. Prior epidemiological investigations have showed that the frequency of menopausal symptoms vary greatly among ladies of varied ethnic backgrounds^12^. For instance, White women reported a significant prevalence of vasomotor symptoms, such as hot flashes and night sweats. In contrast, Asian women generally had physical and somatic symptoms, including headaches, MSKD-like body or joint pains, and sleeping difficulties^13,14^. According to statistics, the occurrence of musculoskeletal diseases (MSDs) among women who have gone through menopause is 27.3% in Nepal and 53.3% in northern India^15^. Among post-menopausal women in Bangladesh, the rate of muscular, skeletal, or muscle and joint discomfort or pain is substantially higher, at 76.20% ^16^. Multiple factors may influence the nutritional health and eating habits of elderly adults. Notwithstanding these adjustments, adhering to a balanced diet and adopting a health-conscious way of life may alone boost the nutritional status and overall well-being of the older demographic^17^. The worldwide number of women who have experienced menopause is growing. Women aged 50 and beyond formed 26% of the total female population globally in 2021. This number grew from 22% a decade ago^18^. Postmenopausal women not only have an increase, but also have a greater prevalence of MSkD^19^.

The purpose of this research was to explore the association between consistent dietary patterns and musculoskeletal disorders (MSKDs) in postmenopausal women in Bangladesh.

## Method

### Study design and participants

We performed descriptive cross-sectional research to assess the correlation between food habits and the health of post-menopausal women with prevalent musculoskeletal illnesses. The study focused on post-menopausal women who received treatment at the Centre for the Rehabilitation of the Paralyzed (CRP), specifically the Musculo-Skeletal Department, located in Savar, CRP-Dhaka Mirpur Centre, and Unique Pain and Paralysis Centre (UPPC), Dhaka Mirpur, during the period from December 2022 to December 2023. We enlisted female individuals ranging in age from 42 to over 70 years, with a total participant count of 576. We used face-to-face interviews as a means of collecting data on demographic factors, medical history, menopausal state, and symptoms (specifically in women), using a standardized questionnaire. The individuals who were excluded from the study were those who met the following criteria: being under the age of 40; women with chronic illnesses such as cancer or HIV/AIDS; women who experienced surgically induced menopause, such as through a hysterectomy; women with polycystic ovary syndrome; and women with neurological disorders, communication disorders, or hearing impairment. Written informed permission was obtained from all participants, and the research received approval from the Ethical Review Committee of the Centre for the Rehabilitation of the Paralyzed (CRP) (Ref: CRP-R&E-0401-0411).

### Measurements

Musculoskeletal problems were characterized using the Bangla version of the Nordic MSK Questionnaire (NMQ), which has been validated and widely used in Bangladesh (Standardized Nordic MSK Questionnaire (NMQ) (1987)). This study found that the specificity ranged from 71% to 88% and the sensitivity ranged from 66% to 92% when compared to earlier MSK surveys.20 The NMQ is divided into three sections: part one covers health-related topics, part two offers data on reproductive factors, and part three asks about musculoskeletal issues. For our research project, we specifically used section 3 of the paper. Using the 24-hour Dietary Consumption Questionnaire, the expert team measured the amount of food consumed by each participant during the day in each category, including carbs, proteins, fats, and miscellaneous items. Grams were used to express the measurements. From the first breakfast meal until the final snack before bed, the total amount of food in each component is calculated. Six daily intervals were set up, and they comprised: Lunch (from 12 to 14 hours), In-between snack (from 14 to 18 hours), Dinner (from 18 to 20 hours), After dinner (after 20 hours), and Breakfast (from the beginning of the day till 09 hours)^21^. The most practical and widely accepted way to measure one’s nutrition intake is by self-reported food intake, which can be measured.

### Assessment of covariates

To evaluate the information provided by all participants, we used a standardized questionnaire presented by an interviewer. The following factors were utilized in the analysis: age, place of residence, degree of education, work status, marital status, average monthly family income, alcohol use, smoking behaviors, regular physical activity, body mass index (BMI) categories, and the Visual Analogue Scale. The BMI is used to distinguish between excessive body fat and has a strong link with a variety of metabolic and disease risk factors^22^. We classified it into four categories by dividing the weight, measured in kilograms, by the square of the height, recorded in meters^23^. BMI categories include underweight (BMI <18.5 kg/m2), normal weight (18.5 kg/m2 ≤ BMI < 24.9 kg/m2), overweight (25 kg/m2 ≤ BMI <29.9 kg/m2), and obese (BMI ≥ 30 kg/m2). Pain is measured using visual analogue scales (VAS), a psychometric instrument^24^. It is a continuous scale of around 10cm in length, anchored by two spoken descriptions, with 0 representing “no pain” and 10 representing “worst imaginable pain.” The proposed cut points for the VAS are as follows: 0 = no pain, 10 - 44mm = mild pain, 45 - 74mm = moderate pain, and 75 - 100 mm = severe pain.

### Statistical analysis

The chi-square test or Mann-Whitney U test were used to compare the demographic features, which were displayed as numbers and frequency distributions for categorical variables or median and interquartile range for continuous data. We estimated mean NMQ scores adjusted for the 24-hour Dietary Intake Questionnaire using generalized correlation models. We also accounted for study locations, domicile, education, employment status, marital status, and average monthly household income. SPSS version ^26^ was used for the analyses. Significant differences were declared at the 0.05 level, and all P values are two-sided.

## RESULTS

### Characteristics of study participants

There were a total of 576 competitors. The largest group of participants, 140 (24.3%), ranged in age from 47 to 70. Out of these people, 49 (16.9%) lived in cities, while 91 (31.8%) lived in rural areas. Of the total, 290 people (50.3%) came from cities, while 286 people (49.7%) came from the countryside. The participants’ educational attainment was divided into the following categories: secondary (198, 34.4%), primary (137, 23.8%), upper secondary (127, 22%), illiterate (56, 9.7%), graduate (47, 8.2%), and post-graduate (11, 1.9%). Of the total participants, 492 (85.4%) were married. The urban region had 230 people, or 79.3% of the total population, while the rural area had 262, or 91%. The study included 525 adults (91.1%) who were unemployed and 51 participants (8.9%) who worked. Yes. The Nuclear Family group had the most participants (369), accounting for 64.1% of the total. The Joint Family category had 196 members, or 34% of the total. The Alone category had the fewest participants, with 11 people making up 1.9% of the total. None of the individuals have had a menstrual cycle in the last three months. Figure 1 shows that 8 participants were classified as underweight, 293 as normal weight, 222 as overweight, and 53 as obese.

**FIG 1.**
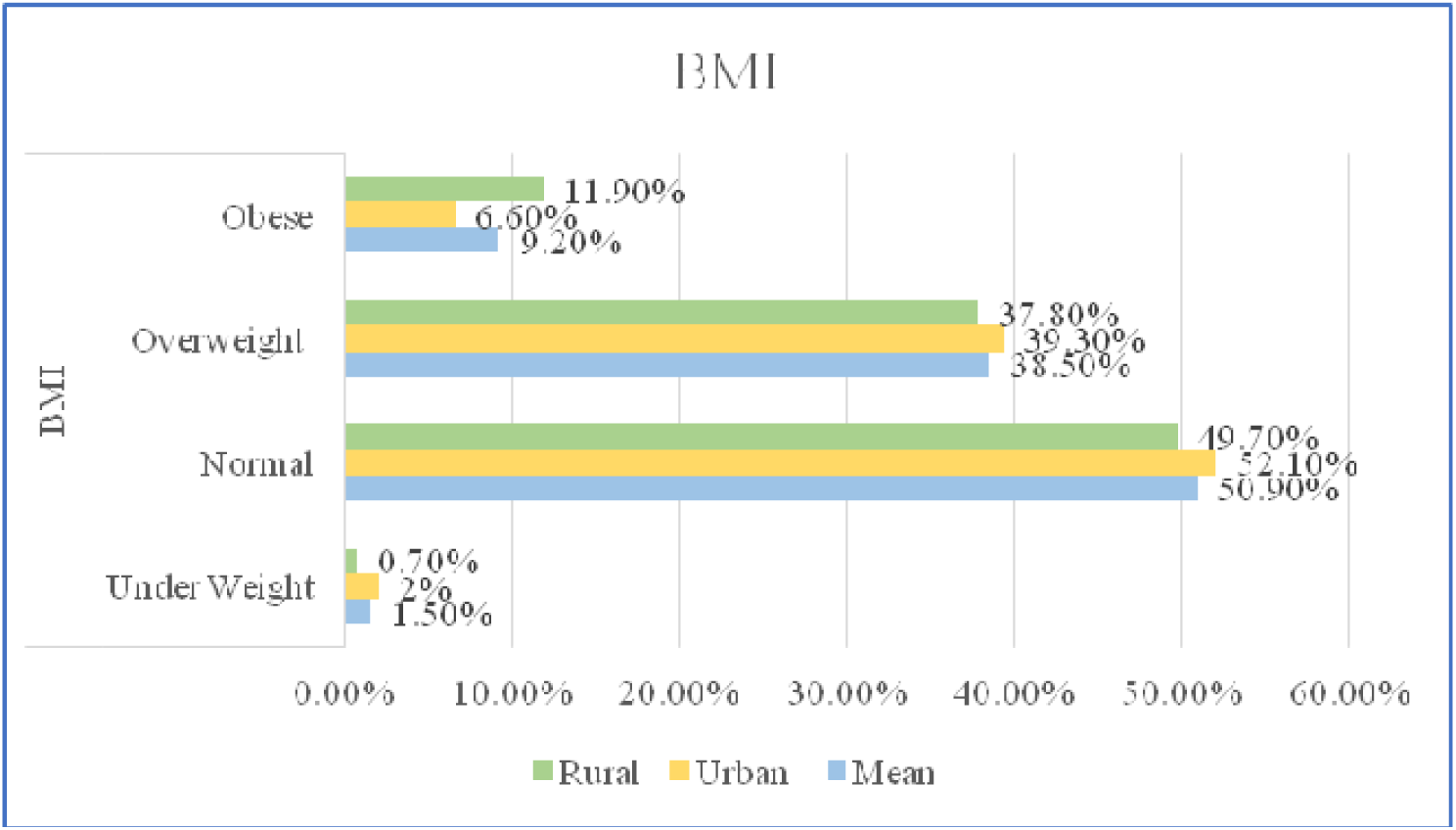
BMI of the participants

Musculoskeletal problems are described in Table 2. There were 576 participants in the study, all of whom had felt pain or discomfort. Of them, 286 were from the rural area and 290 were from the metropolitan area. Overall neck pain (13.8%), shoulder pain (9.4%), upper back pain (5%), elbow pain (3.8%), wrist or hand pain (2.3%), low back pain (33.2%), hip or thigh pain (4.3%), knee pain (23.5%), and ankle or foot pain (4.7%) were the most common types of pain among the participants. Other issues that the people displayed were fecal incontinence (8.1%), pelvic organ prolapses (58 instances, 10.1%), and urine incontinence (100 cases, 17.4%). Asymmetrical pain was reported by 326 participants (56.6%), while symmetrical pain was reported by 250 participants (43.4%). 132 (22.9%) was the maximum amount of discomfort felt in the morning, and as the day wore on, it rose to 192 (33.3%). The discomfort dropped to 29 (5%) in the evening but increased to 223 (38.7%) at night. Participants reported 60 cases (10.4%) of intermittent discomfort, 186 cases (32.3%) of chronic pain, and 330 cases (57.3%) of recurrent pain. Swelling occurred in 143 cases (15.5%), stiffness in 173 cases (18.7%), paresthesia in 170 cases (18.4%), tingling in 274 cases (29.7%), numbness in 75 cases (8.1%), cramping in 79 cases (8.5%), and contracture in 10 cases (1.1%) for every symptom that each participant displayed. Of all participants, 164 (28.5%) said that pain made it hard for them to fall asleep. Furthermore, 125 (21.7%) reported having this problem once a week, 211 (36.6%) reported having it two to three times a week, 52 (9%) reported having it four to five times a week, and 24 (4.2%) reported having it more than five times a week. Thirty-one (5.4%) of the subjects did not experience mood fluctuations associated with musculoskeletal issues, depression, gloomy mood, or lack of vigor. 33 (5.7%) had severe symptoms, 1 (0.2%) had really severe symptoms, 284 (49.3%) had light symptoms, and 227 (39.4%) had moderate symptoms. The current problem was experienced by 141 participants (24.5%) during the first three months, 165 participants (28.6%) during the fourth month, 95 participants (16.5%) during the fifth month, 51 individuals (8.9%) during the sixth month, and 124 participants (21.5%) during the seventh month. A pain rating of 6.11±1.321 was recorded on the VAS scale; the urban area reported a pain rating of 6.44±1.338, while the rural area reported a pain rating of 5.77±1.213 out of 10.

**TABLE 1.**
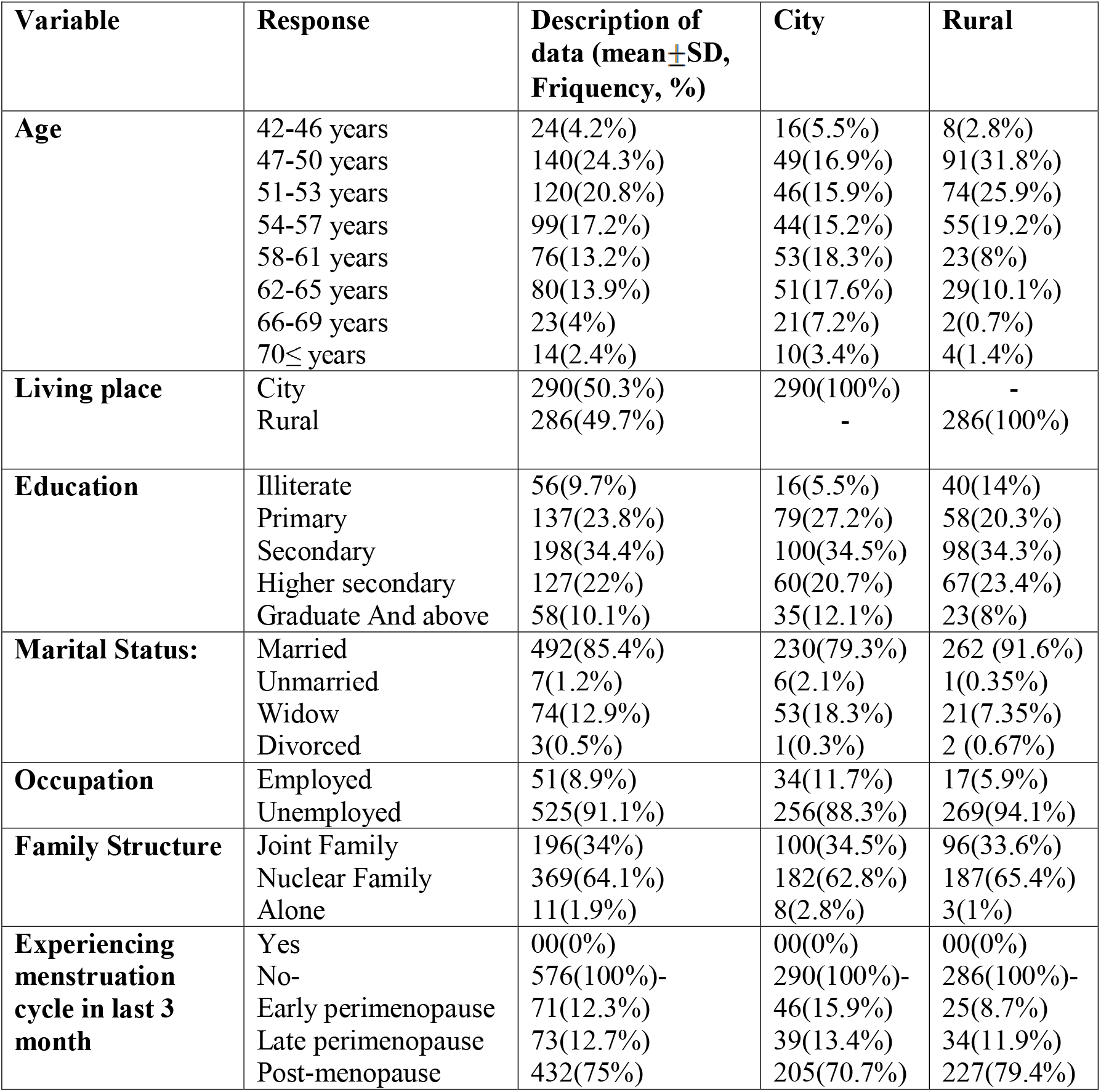
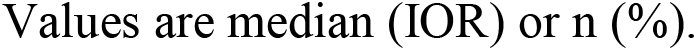
Characteristics of participants on the basis of post-menopausal status.

| Variable | Response | Description of data (mean±SD, Friquency, %) | City | Rural |
| --- | --- | --- | --- | --- |
| **Age** | 42-46 years | 24(4.2%) | 16(5.5%) | 8(2.8%) |
|  | 47-50 years | 140(24.3%) | 49(16.9%) | 91(31.8%) |
|  | 51-53 years | 120(20.8%) | 46(15.9%) | 74(25.9%) |
|  | 54-57 years | 99(17.2%) | 44(15.2%) | 55(19.2%) |
|  | 58-61 years | 76(13.2%) | 53(18.3%) | 23(8%) |
|  | 62-65 years | 80(13.9%) | 51(17.6%) | 29(10.1%) |
|  | 66-69 years | 23(4%) | 21(7.2%) | 2(0.7%) |
| | 70≤ years | 14(2.4%) | 10(3.4%) | 4(1.4%) |
| **Living place** | City | 290(50.3%) | 290(100%) | - |
|  | Rural | 286(49.7%) | - | 286(100%) |
| **Education** | Illiterate | 56(9.7%) | 16(5.5%) | 40(14%) |
|  | Primary | 137(23.8%) | 79(27.2%) | 58(20.3%) |
|  | Secondary | 198(34.4%) | 100(34.5%) | 98(34.3%) |
|  | Higher secondary | 127(22%) | 60(20.7%) | 67(23.4%) |
|  | Graduate And above | 58(10.1%) | 35(12.1%) | 23(8%) |
| **Marital Status:** | Married | 492(85.4%) | 230(79.3%) | 262 (91.6%) |
|  | Unmarried | 7(1.2%) | 6(2.1%) | 1(0.35%) |
|  | Widow | 74(12.9%) | 53(18.3%) | 21(7.35%) |
|  | Divorced | 3(0.5%) | 1(0.3%) | 2 (0.67%) |
| **Occupation** | Employed | 51(8.9%) | 34(11.7%) | 17(5.9%) |
|  | Unemployed | 525(91.1%) | 256(88.3%) | 269(94.1%) |
| **Family Structure** | Joint Family | 196(34%) | 100(34.5%) | 96(33.6%) |
|  | Nuclear Family | 369(64.1%) | 182(62.8%) | 187(65.4%) |
|  | Alone | 11(1.9%) | 8(2.8%) | 3(1%) |
| **Experiencing menstruation cycle in last 3 month** | Yes | 00(0%) | 00(0%) | 00(0%) |
|  | No- | 576(100%)- | 290(100%)- | 286(100%)- |
|  | Early perimenopause | 71(12.3%) | 46(15.9%) | 25(8.7%) |
|  | Late perimenopause | 73(12.7%) | 39(13.4%) | 34(11.9%) |
|  | Post-menopause | 432(75%) | 205(70.7%) | 227(79.4%) |

Values are median (IOR) or n (%).

**TABLE 2.** Musculoskeletal complain information.

| Variable | Response | Description of data (mean±SD, Frequency, %) | City | Rural |
| --- | --- | --- | --- | --- |
| **Feeling discomfort or pain on body last 12 months** | No<br>Yes | 00 (0%)<br>576(100%) | 00 (0%)<br>290(100%) | 00 (0%)<br>286(100%) |
| **body part is affected** | Neck<br>Shoulder<br>Upper back<br>Elbow<br>Wrist/hand<br>Low back<br>Hip/thigh<br>Knee<br>Ankle/feet | 110(13.8%)<br>75(9.4%)<br>40(5%)<br>30(3.8%)<br>18(2.3%)<br>264(33.2%)<br>34(4.3%)<br>187(23.5%)<br>37(4.7%) | 68(16.4%)<br>42(10.1%)<br>20(4.8%)<br>15(3.6%)<br>8(1.4%)<br>129(31.1%)<br>15(3.6%)<br>98(23.6%)<br>22(5.3%) | 42(11.2%)<br>33(8.3%)<br>20(5.3%)<br>15(4%)<br>10(2.7%)<br>135(36%)<br>19(5.1%)<br>89(23.7%)<br>15(4%) |
| **suffering from others condition** | Urinary incontinence<br>Pelvic organ prolapses<br>Fecal incontinence<br>none | 100(17.4%)<br>58(10.1%)<br>8(1.4%)<br>410(71.2%) | 62(21.4%)<br>21(7.2%)<br>4(1.4%)<br>203(70%) | 38(13.3%)<br>37(12.9%)<br>4(1.4%)<br>207(72.4%) |
| **Pattern of pain** | Symmetrical<br>Asymmetrical | 250(43.4%)<br>326(56.6%) | 146(50.3%)<br>144(49.7%) | 104(36.3%)<br>182(63.6%) |
| **pick time of pain.** | Morning<br>Day<br>Evening<br>Night | 132(22.9%)<br>192(33.3%)<br>29(5%)<br>223(38.7%) | 93(32.1%)<br>40(13.8%)<br>22(7.6%)<br>135(46.6%) | 39(13.6%)<br>152(53.1%)<br>7(2.4%)<br>88(30.8%) |
| **Type of pain** | Continuous<br>Intermittent<br>Occasional | 186(32.3%)<br>330(57.3%)<br>60(10.4%) | 106(36.6%)<br>138(47.6%)<br>46(15.9%) | 80(28%)<br>192(67.1%)<br>14(4.9%) |
| **Post menopausal symptoms** | Swelling<br>Stiffness<br>Paresthesia<br>Tingling<br>Numbness<br>Cramping<br>Contracture | 143(15.5%)<br>173(18.7%)<br>170(18.4%)<br>274(29.7%)<br>75(8.1%)<br>79(8.5%)<br>10(1.1%) | 83(18.9%)<br>76(17.3%)<br>78(17.7%)<br>113(25.7%)<br>37(8.4%)<br>51(11.6%)<br>2(0.5%) | 60(12.4%)<br>97(20%)<br>92(19%)<br>161(33.3%)<br>38(7.9%)<br>28(5.8%)<br>8(1.7%) |
| **Trouble in sleeping because of pain** | Not at all | 164(28.5%) | 51(17.6%) | 113(39.5%) |
|  | 1 times/weeks | 125(21.7%) | 58(20%) | 67(23.4%) |
|  | 2-3 time/weeks | 211(36.6%) | 127(43.8%) | 84(29.4%) |
|  | 4-5 times/weeks | 52(9%) | 38(13.1%) | 14(4.9%) |
|  | >5 times/week | 24(4.2%) | 16(5.5%) | 8(2.8%) |
| **Experienced depressive mood, sad, lack of thrive, mood swings due to MSK problem** | Not at all | 31(5.4%) | 21(7.2%) | 10(3.5%) |
|  | Mild | 284(49.3%) | 118(40.7%) | 166(58%) |
|  | Moderate | 227(39.4%) | 132(45.5%) | 95(33.2%) |
|  | Severe | 33(5.7%) | 18(6.2%) | 15(5.2%) |
|  | Very severe | 1(0.2%) | 1(0.3%) | 0(0%) |
| **How long suffer from current problem.** | 1-3 month | 141(24.5%) | 50(17.2%) | 91(31.8%) |
|  | 4-6 month | 165(28.6%) | 84(29%) | 81(28.3%) |
|  | 7-9 month | 95(16.5%) | 38(13.1%) | 57(19.9%) |
|  | 10-12 month | 51(8.9%) | 32(11%) | 19(6.6%) |
|  | >more than year | 124(21.5%) | 86(29.7%) | 38(13.3%) |
| **Rate your pain** | **in VAS scale** | 6.11±1.321 | 6.44±1.338 | 5.77±1.213 |

Values are median (IOR) or n (%).

Figure 2 depicts the nutritional intake of each participant throughout a 24-hour period. The average daily protein intake was 224.73 grams, with urban dwellers eating 233.65 grams and rural residents eating 215.68 grams. An adult’s daily recommended intake should range between 50 and 175 grams. The average daily carbohydrate consumption for urban and rural residents was 641.20 grams, 573.38 grams, and 709.97 grams, respectively. The recommended daily consumption for an adult is between 225 and 325 grams. The average fat intake was 31.72 grams, with the city population taking 35.82 grams and the village population ingesting 27.56 grams. An adult’s daily intake should range from 44 to 78 grams. City dwellers consumed 317.67 grams of vegetables, but rural dwellers consumed 278.36 grams. In terms of fruits, city dwellers ate 103.85 grams, whereas rural residents ate 87.27. Overall, veggies accounted for 298.15 grams, while fruits accounted for 95.62 grams.

**FIG 2.**
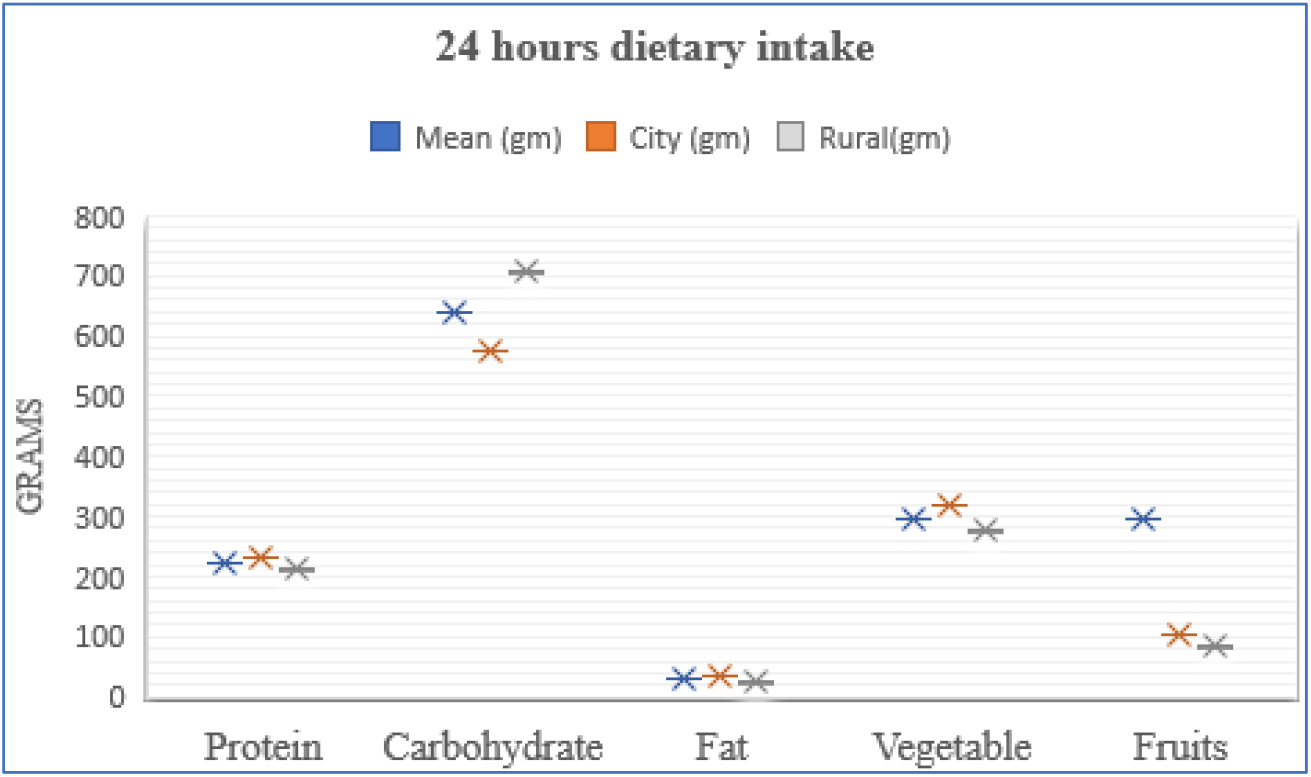
**24 hours dietary intake**

### Proportion of chronic musculoskeletal pain and dietary elements

Table 3 reveals a significant protein association (0.046*) with upper back pain. Carbohydrate was associated with LBP (0.005*), hip pain (0.008*), and knee pain (0.002*), while fat was associated with shoulder pain (0.007*), wrist pain (0.002*), hip pain (0.044*), knee pain (0.002*), and ankle pain (0.010*).

**TABLE 3.**
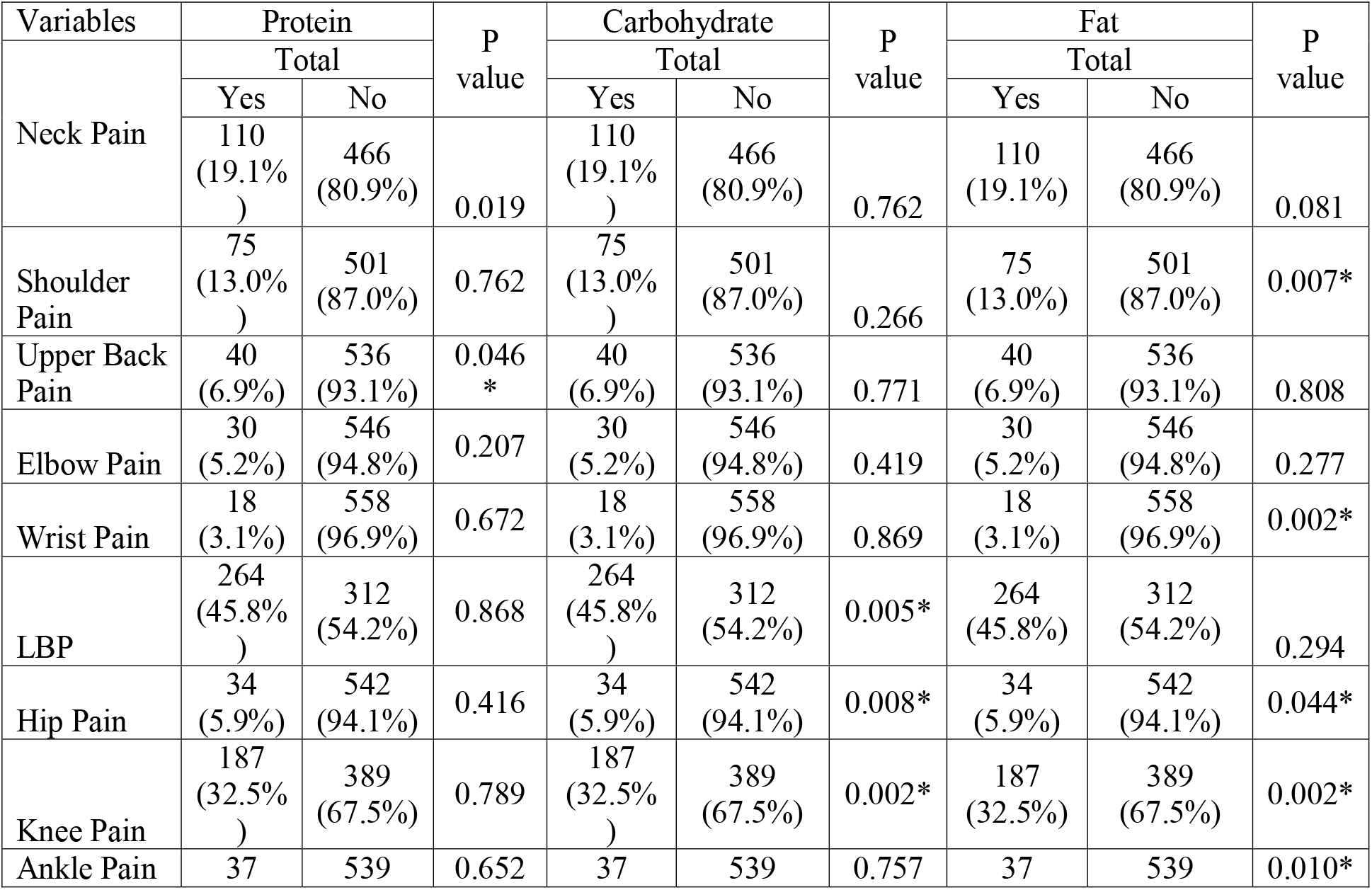
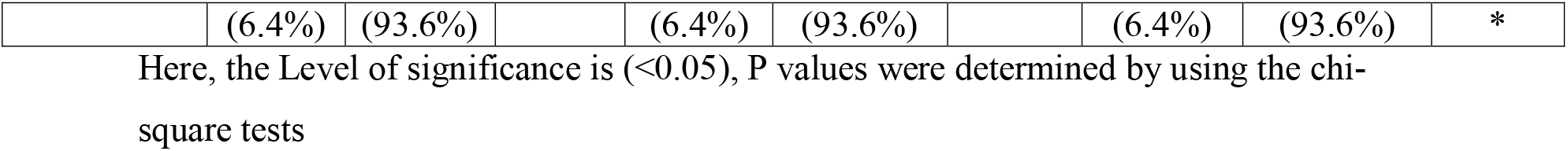
Chronic musculoskeletal pain and dietary elements.

| Variables | Protein |  | P<br>value | Carbohydrate |  | P<br>value | Fat |  | P<br>value |
| --- | --- | --- | --- | --- | --- | --- | --- | --- | --- |
|  | Total |  |  | Total |  |  | Total |  |  |
|  | Yes | No |  | Yes | No |  | Yes | No |  |
| Neck Pain | 110<br>(19.1%<br>) | 466<br>(80.9%) | 0.019 | 110<br>(19.1%<br>) | 466<br>(80.9%) | 0.762 | 110<br>(19.1%) | 466<br>(80.9%) | 0.081 |
| Shoulder Pain | 75<br>(13.0%<br>) | 501<br>(87.0%) | 0.762 | 75<br>(13.0%<br>) | 501<br>(87.0%) | 0.266 | 75<br>(13.0%) | 501<br>(87.0%) | 0.007* |
| Upper Back Pain | 40<br>(6.9%) | 536<br>(93.1%) | 0.046<br>* | 40<br>(6.9%) | 536<br>(93.1%) | 0.771 | 40<br>(6.9%) | 536<br>(93.1%) | 0.808 |
| Elbow Pain | 30<br>(5.2%) | 546<br>(94.8%) | 0.207 | 30<br>(5.2%) | 546<br>(94.8%) | 0.419 | 30<br>(5.2%) | 546<br>(94.8%) | 0.277 |
| Wrist Pain | 18<br>(3.1%) | 558<br>(96.9%) | 0.672 | 18<br>(3.1%) | 558<br>(96.9%) | 0.869 | 18<br>(3.1%) | 558<br>(96.9%) | 0.002* |
| LBP | 264<br>(45.8%<br>) | 312<br>(54.2%) | 0.868 | 264<br>(45.8%<br>) | 312<br>(54.2%) | 0.005* | 264<br>(45.8%) | 312<br>(54.2%) | 0.294 |
| Hip Pain | 34<br>(5.9%) | 542<br>(94.1%) | 0.416 | 34<br>(5.9%) | 542<br>(94.1%) | 0.008* | 34<br>(5.9%) | 542<br>(94.1%) | 0.044* |
| Knee Pain | 187<br>(32.5%<br>) | 389<br>(67.5%) | 0.789 | 187<br>(32.5%<br>) | 389<br>(67.5%) | 0.002* | 187<br>(32.5%) | 389<br>(67.5%) | 0.002* |
| Ankle Pain | 37 | 539 | 0.652 | 37 | 539 | 0.757 | 37 | 539 | 0.010* |
|  | (6.4%) | (93.6%) |  | (6.4%) | (93.6%) |  | (6.4%) | (93.6%) | * |

Here, the Level of significance is ( $<0.05$ ), P values were determined by using the chi-square tests

### Association between patient’s rated acute pain (pain in last week) and dietary elements by Pearson’s correlation

Table 4 revealed that there is no statistically significant association between acute pain (pain in the last week) and protein (r= −0.015; p= 0.713), fat (r= 0.075; p= 0.072), vegetable (r= 0.045; p= 0.278), fruits (r= −0.021; p= 0.615), but carbohydrate has an association with acute pain (r= −0.104; p= 0.012).

**TABLE 4.**
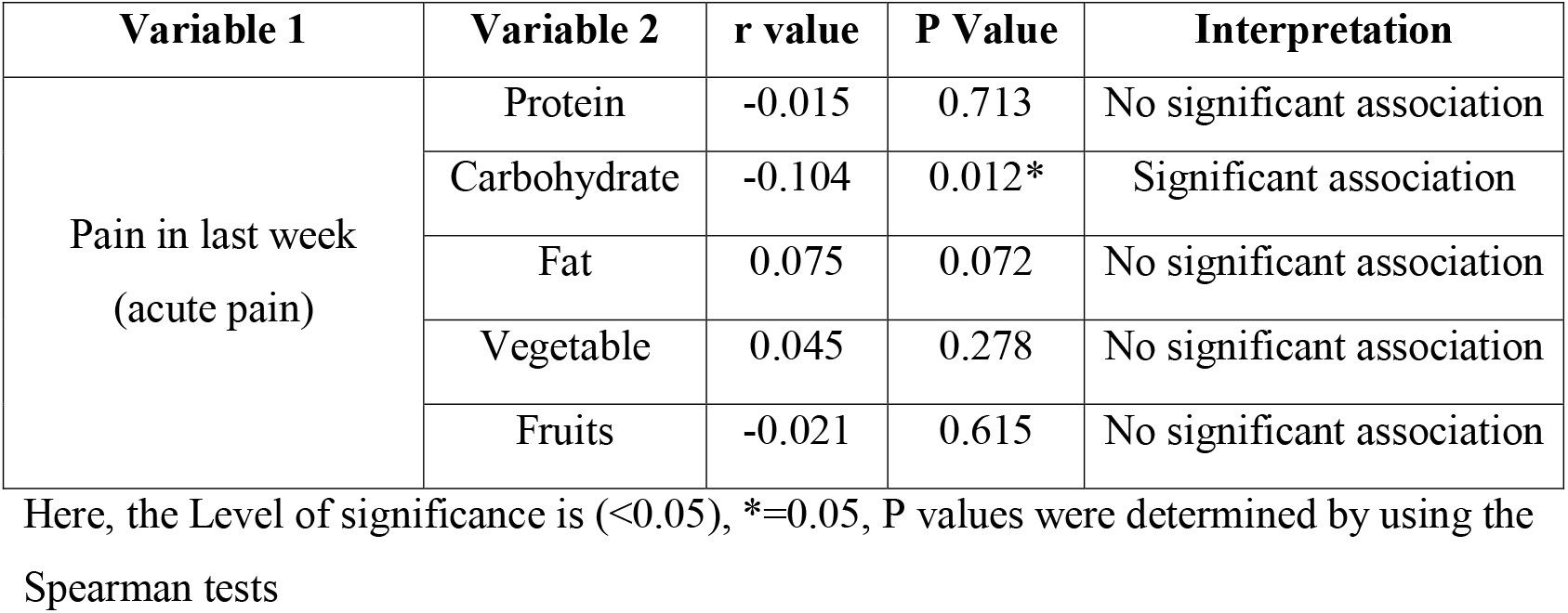
Cross tabulation between acute pain (pain in last week) and dietary elements.

| Variable 1 | Variable 2 | r value | P Value | Interpretation |
| --- | --- | --- | --- | --- |
| Pain in last week<br>(acute pain) | Protein | -0.015 | 0.713 | No significant association |
|  | Carbohydrate | -0.104 | 0.012* | Significant association |
|  | Fat | 0.075 | 0.072 | No significant association |
|  | Vegetable | 0.045 | 0.278 | No significant association |
|  | Fruits | -0.021 | 0.615 | No significant association |

Here, the Level of significance is ( $<0.05$ ), $*=0.05$ , P values were determined by using the Spearman tests

## Discussion

The participants in this study ranged in age from 47 to 50, with the majority of them (140 out of 586) falling within this age range, accounting for 24.3% of the total. The next highest age group was 51 to 53 years old, with 120 people (20.8%). A study confirmed the study’s findings by stating that menopause normally occurs between the ages of 45 and 55. The UK menopause guideline (2012) provided similar numbers, suggesting that the age range for menopause onset is typically 45 to 55 years, with the average age of the last menstrual cycle being 52 years^1^.

According to the Kuppusamy socioeconomic level scale, a substantial majority of those who reported musculoskeletal disorders or complaints were from the upper middle class-II category (16-25), which included 280 respondents (48.6%). The lower middle class-III category (11-15) had the second greatest number of responders (204, 35.4%). Another study found that people who live in less affluent locations are more likely to experience severe musculoskeletal pain than those who live in more rich areas^25^.

The current study employs the Nordic Musculoskeletal Questionnaire to identify the exact joints or body locations in which menopausal women experience discomfort or pain throughout their daily activities. According to the findings, the majority of respondents (264, 33.2%) experience lower back issues. On average, rural women experience this condition in 135 cases (36%), while city women experience it in 129 cases (31.1%). Knee soreness is the second most common complaint, with 187 cases (23.5%). Urban women reported a somewhat higher rate (23.6%) than rural women (89 occurrences). As a result, a total of 110 women (13.8%), 68 of whom lived in the city, experienced neck stiffness. This was followed by 75.4% shoulder pain, 5% upper back pain, 3.7% ankle pain, and 4.3% hip pain. This investigation confirms that menopausal musculoskeletal complaints are more severe on a somatic-vegetative level, particularly in the neck, elbow, shoulder, and knees^26^. Musculoskeletal pain is most frequently felt in the lower extremities and back^27^. A study in Bangladesh discovered that the joints, including the neck, shoulder, and lower back, are the most commonly impacted by musculoskeletal discomfort after menopause28. According to recent research conducted in Bangladesh, the most common body locations affected after menopause are the neck, shoulder, lower back, and knee^29^. Musculoskeletal issues are also associated with symptoms such as urine incontinence (17.4%), pelvic organ prolapse (10.1%), and fecal incontinence (1.4%). According to one study, women’s bodies change in a variety of ways after menopause. These changes include genitourinary issues like discomfort, dryness, and persistent urinary tract infections (UTIs). The connective tissue that supports the pelvic organs also weakens. During this time, women have urogenital atrophy as well as sexual issues.

The current study also found some significant links between carbohydrate consumption and chronic musculoskeletal pain, including lower back pain (p = 0.005), hip pain (p = 0.008), and knee pain (p = 0.002). Similar findings were seen when there was a significant link between fat consumption and shoulder pain (0.007), wrist pain (0.002), hip pain (0.044), knee pain (0.002), and ankle pain (0.010). Furthermore, there was a significant association between protein intake and upper back pain (0.046). In contrast, a study discovered a link between chronic physical and psychological complaints in postmenopausal women, high-quality carbohydrate consumption, and successful dietary adjustments ^30^.

Furthermore, studies have found no link between acute discomfort (pain experienced within the recent week) and dietary variables. Contrary research disputes the notion that eating more fruits and vegetables, as well as specific vitamins and minerals, might effectively reduce acute pain^31^. As a result, research reveals that there is a substantial relationship between the severity of menopausal symptoms and the quality of nutritional and food consumption, as well as dietary habits, in postmenopausal women^32^.

This study is particularly robust because it assesses menopausal symptoms using a validated measure and thoroughly accounts for any variables that may impact the findings. The study included people who lived in both urban and rural environments. The study includes some notable limitations. Because of the cross-sectional research design, it is impossible to demonstrate a causal link between musculoskeletal problems and eating habits. The second sample size is relatively small. Finally, despite our recommendation against using hormones, we were unable to assess their impact on postmenopausal women due to a lack of adequate data.

## Conclusion

In conclusion, our study demonstrated very little connection between post-menopausal women’s normal eating habits and acute musculoskeletal pain. Nonetheless, a significant correlation was observed with certain long-term musculoskeletal conditions. To validate our findings and explore the potential mechanisms linking postmenopausal women with musculoskeletal disorders (MSKDs) to their typical eating patterns, more study is needed.

## Data Availability

All data produced in the present work are contained in the manuscript

## REFERENCES

1. Ahmed K, Jahan P, Nadia I, Ahmed F. Assessment of menopausal symptoms among early and late menopausal midlife Bangladeshi women and their impact on the quality of life. Journal of menopausal medicine 2016; 22:39–46.

2. Who SG. Research on the Menopause in the 1990’s: A Report of the WHO Scientific Group. Geneva: World Health Organization 1996; 1:07.

3. Abedzadeh-Kalahroudi M, Taebi M, Sadat Z, Saberi F, Karimian Z. Prevalence and severity of menopausal symptoms and related factors among women 40-60 years in Kashan, Iran. Nurs Midwifery Stud 2012; 1:88–93.

4. Yasmin N, Sultana S, Habib SA, Khatun K. Intervention approach to the menopausal women in rural Bangladesh. Bangladesh Medical Journal 2009; 38:9–14.

5. Wolf JM, Cannada L, Van Heest AE, O’Connor MI, Ladd AL. Male and female differences in musculoskeletal disease. JAAOS-Journal of the American Academy of Orthopaedic Surgeons 2015; 23:339–47.

6. Van Dijk GM, Kavousi M, Troup J, Franco OH. Health issues for menopausal women: the top 11 conditions have common solutions. Maturitas 2015; 80:24–30.

7. Lim SS, Vos T, Flaxman AD, Danaei G, Shibuya K, Adair-Rohani H, AlMazroa MA, Amann M, Anderson HR, Andrews KG, Aryee M. A comparative risk assessment of burden of disease and injury attributable to 67 risk factors and risk factor clusters in 21 regions, 1990–2010: a systematic analysis for the Global Burden of Disease Study 2010. The lancet 2012; 380:2224–60.

8. Khadilkar SS. Musculoskeletal disorders and menopause. The Journal of Obstetrics and Gynecology of India 2019; 69:99–103.

9. Ranasinghe C, Shettigar PG, Garg M. Dietary intake, physical activity and body mass index among postmenopausal women. Journal of mid-life health 2017; 8:163–9.

10. Crandall CJ, Mehta JM, Manson JE. Management of menopausal symptoms: a review. Jama 2023; 329:405–20.

11. Burkard T, Moser M, Rauch M, Jick SS, Meier CR. Utilization pattern of hormone therapy in UK general practice between 1996 and 2015: a descriptive study. Menopause 2019; 26:741–9.

12. Monteleone P, Mascagni G, Giannini A, Genazzani AR, Simoncini T. Symptoms of menopause—global prevalence, physiology and implications. Nature Reviews Endocrinology 2018; 14:199–215.

13. Islam MR, Gartoulla P, Bell RJ, Fradkin P, Davis SR. Prevalence of menopausal symptoms in Asian midlife women: a systematic review. Climacteric 2015; 18:157–76.

14. Baber RJ. East is east and West is west: perspectives on the menopause in Asia and The West. Climacteric 2014;17: 23–8.

15. Acharya D, Gautam S, Neupane N, Kaphle HP, Singh JK. Health problems of women above forty years of age in rupandehi district of Nepal. Int J Health Sci Res 201; 3:29–36.

16. Rahman S, Salehin F, Iqbal A. Menopausal symptoms assessment among middle age women in Kushtia, Bangladesh. BMC research notes 2011; 4:1–4.

17. Bakker R. Sensory loss, dementia, and environments. Generations 2003; 27:46–51.

18. United Nations, Department of Economic and Social Affairs. World population Prospects 2021. Available at: https://population.un.org/wpp/. Accessed August 01, 2024.

19. Dugan SA, Powell LH, Kravitz HM, Rose SA, Karavolos K, Luborsky J. Musculoskeletal pain and menopausal status. The Clinical journal of pain 2006; 22:325–31.

20. Ohlsson K, Attewell RG, Johnsson B, Ahlm A, Skerfving S. An assessment of neck and upper extremity disorders by questionnaire and clinical examination. Ergonomics 1994; 37:891–7.

21. DeBiasse MA, Bowen DJ, Quatromoni PA, Quinn E, Quintiliani LM. Feasibility and acceptability of dietary intake assessment via 24-hour recall and food frequency questionnaire among women with low socioeconomic status. Journal of the Academy of Nutrition and Dietetics 2018; 118:301–7.

22. Farias Júnior JC, Konrad LM, Rabacow FM, Araújo VC. Sensitivity and specificity of criteria for classifying body mass index in adolescents. Revista de Saúde Publica 2009; 43:53–9.

23. Wang Y, Mi J, Shan XY, Wang QJ, Ge KY. Is China facing an obesity epidemic and the consequences? The trends in obesity and chronic disease in China. International journal of obesity 2007; 31:177–88.

24. Klimek L, Bergmann KC, Biedermann T, Bousquet J, Hellings P, Jung K, Merk H, Olze H, Schlenter W, Stock P, Ring J. Visual analogue scales (VAS): Measuring instruments for the documentation of symptoms and therapy monitoring in cases of allergic rhinitis in everyday health care: Position Paper of the German Society of Allergology (AeDA) and the German Society of Allergy and Clinical Immunology (DGAKI), ENT Section, in collaboration with the working group on Clinical Immunology, Allergology and Environmental Medicine of the German Society of Otorhinolaryngology, Head and Neck Surgery (DGHNOKHC). Allergo journal international 2017; 26:16–24.

25. Brekke M, Hjortdahl P, Kvien TK. Severity of musculoskeletal pain: relations to socioeconomic inequality. Social science & medicine 2002; 54:221–8.

26. Santo JE, Lavilla□Lerma ML, del Carmen Carcelén□Fraile M, de Loureiro NE, Brandão□Loureiro V, Alzar□Teruel M, Ortiz□Quesada R. Associations between the severity of menopausal symptoms and musculoskeletal pain in postmenopausal Portuguese women. International Journal of Gynecology & Obstetrics 2024; 165:138–47.

27. Ogwumike OO, Adeniyi AF, Orogbemi OO. Musculoskeletal pain among postmenopausal women in Nigeria: Association with overall and central obesity. Hong Kong Physiotherapy Journal 2016; 34:41–6.

28. Biswas T, Uddin MJ, Mamun AA, Pervin S P, Garnett S. Increasing prevalence of overweight and obesity in Bangladeshi women of reproductive age: findings from 2004 to 2014. PloS one 2017;12: e0181080.

29. Akter N, Haseen F, Hasan M, Haney U, Bristy S, Islam SS. Nutritional Knowledge and Dietary Diversity of Post-menopausal Women in Rural Areas of Bangladesh. Journal of Health and Medical Science 2023; 6:22–29.

30. Mohsenian S, Shabbidar S, Siassi F, Qorbani M, Khosravi S, Abshirini M, Aslani Z, Sotoudeh G. Carbohydrate quality index: Its relationship to menopausal symptoms in postmenopausal women. Maturitas 202; 150:42–8.

31. Qiu R, Cao WT, Tian HY, He J, Chen GD, Chen YM. Greater intake of fruit and vegetables is associated with greater bone mineral density and lower osteoporosis risk in middle-aged and elderly adults. PLoS One 2017; 12: e0168906.

32. Soleymani M, Siassi F, Qorbani M, Khosravi S, Aslany Z, Abshirini M, Zolfaghari G, Sotoudeh G. Dietary patterns and their association with menopausal symptoms: a cross-sectional study. Menopause 2019; 26:365–72.

